# A new method using rapid Nanopore metagenomic cell-free DNA sequencing to diagnose bloodstream infections: a prospective observational study

**DOI:** 10.1101/2024.05.09.24307053

**Authors:** Morten Eneberg Nielsen, Kirstine Kobberøe Søgaard, Søren Michael Karst, Anne Lund Krarup, Hans Linde Nielsen, Mads Albertsen

## Abstract

**Background:** Bloodstream infections (BSIs) remain a major cause of mortality, in part due to many patients developing sepsis or septic shock. To survive sepsis, it is paramount that effective antimicrobial therapy is initiated rapidly to avoid excess mortality, but the current gold-standard to identify the pathogen in BSIs, blood culturing, has great limitations with a long turnaround time and a poor sensitivity. This delay to correct empiric broad-spectrum antimicrobial treatments leads to excess mortality and antimicrobial resistance development.

**Methods:** In this study we developed a metagenomic next-generation sequencing (mNGS) assay utilizing the Oxford Nanopore Technologies platform to sequence microbial cell-free DNA from blood plasma. The method was evaluated in a prospective observational clinical study (n=40) in an emergency ward setting, where a study sample was taken from the same venipuncture as a blood culture sample from patients with a suspected BSI.

**Findings:** Nanopore mNGS confirmed all findings in patients with a positive blood culture (n=11), and identified pathogens relevant to the acute infection in an additional 11 patients with a negative blood culture. In an analysis of potential impact on the antibiotic treatment, we found that 59% (n=13) of mNGS positive answers could have impacted the treatment, with five cases of a change from ineffective to effective therapy.

**Interpretation:** This study demonstrates that culture-independent Nanopore mNGS directly on blood plasma could be a feasible alternative to blood culturing for infection diagnostics for patients admitted with a severe infection or sepsis. The method identified a relevant pathogen in patients with a broad range of etiologies including urinary tract infections and lower respiratory tract infections. With a turnaround time of 6 hours the method could provide unprecedented speed and sensitivity in BSI diagnostics.

## Introduction

Bloodstream infections (BSIs) are frequent and associated with a high mortality due to the development of sepsis.^1^ The Global Burden of Disease study,^2^ found that there were 50 million sepsis cases and 11 million sepsis related deaths in 2017, constituting nearly 20% of all global deaths. To treat sepsis, the current international guideline recommends that broad-spectrum empiric antibiotic treatment should be administered ideally within 1 hour from its recognition.^3^ However, the empiric antibiotic treatment does not cover the causative pathogen in 20% of cases, and these patients have increased mortality (data from US and Canada).^4,5^ To facilitate a targeted and effective antibiotic treatment to improve sepsis survival, an accurate and rapid identification of the causative pathogen and its antimicrobial susceptibility is crucial. However, gold-standard blood culturing fails to diagnose >50% of patients with clinical sepsis, possibly due to prior antibiotic treatment or a microbial load below the limit of detection.^6^ Further, blood culturing has a turnaround time of 24-72 hours, which contrasts the urgency to provide effective antibiotic treatment.^7^ In addition to the urgency on a patient level, there is an increased focus on improving diagnostics to facilitate effective antimicrobial stewardship programs.^8,9^

This has prompted the development of novel approaches to BSI diagnostics including multiplex PCR assays, molecular target detection with magnetic resonance, and metagenomic next-generation sequencing (mNGS) of microbial cell-free DNA (cfDNA) in plasma.^10-13^ Particularly mNGS has gained attention due to its high sensitivity and hypothesis-free approach. While cfDNA mNGS has an increased number of relevant detections of pathogens compared to the standard blood culturing, the method has generally been applied on DNA sequencing platforms with long turnaround times that compromise the urgency of diagnosing BSIs. We leveraged the real-time Nanopore sequencing platform to develop a plasma mNGS assay with a short turnaround time and evaluated it in a prospective observational study in an emergency ward setting against gold-standard blood culturing.

## Methods

### Study design

We performed a cross-sectional study in the emergency ward at Aalborg University Hospital, which is the largest hospital in the North Denmark Region with highly specialized functions and a total of 558 beds (catchment population approximately 600,000 inhabitants). Patients were prospectively included upon suspicion of a BSI. All included patients provided written informed consent (see ethics section) and were included in the period from 1 June to 31 December 2022. For included patients, a blood sample (9mL K2EDTA) for mNGS was drawn from the same venipuncture as a blood culture for routine diagnostics. Following inclusion, patient metadata was collected from their electronic medical record, which included medical history and diagnosis, clinical microbiology results, biochemical blood parameters, and antimicrobial therapy. As a negative reference group, blood donor samples were collected from the blood bank at Aalborg University Hospital from 12 donors and handled in parallel with the clinical samples.

### Study population

Study inclusion criteria comprised that the patient was admitted to the emergency ward at Aalborg University Hospital and admitted upon suspicion of a BSI as adjudicated by a requested blood culture. Patients were excluded from the study if they were younger than 18 years old or were unable to provide written informed consent. Inclusion was conducted on selected weekdays from 8AM-4PM.

### Patient subgrouping

Based on the medical records, patients were subdivided into the following groups as adjudicated by a clinical microbiologist (KKS): patients with a positive blood culture (BSI-confirmed); patients with a negative blood culture but a high suspicion of a true BSI (BSI-suspected); patients with a negative blood culture and no suspicion of an infectious disease (BSI-absent); and patients with a negative blood culture and low to medium suspicion of a true BSI (BSI-unlikely). Patients were assigned based on all available microbiology test results (blood culture results, urine culture, respiratory samples etc.) combined with a clinical evaluation of the patients including vital signs (e.g. temperature, pulse, blood pressure, respiration rate), biochemical parameters (e.g. CRP, white blood cell count) and imaging (e.g. chest x-ray, CT abdomen and chest, endoscopies). Patients in groups BSI-confirmed, BSI-suspected, and BSI-absent were selected for analysis.

### Clinical microbiology

All clinical microbiology results were subsequently obtained from patient medical records (wwBakt, Autonik AB, Sweden) and no follow-up analyses were conducted. Clinical microbiology analyses were conducted at the Department of Clinical Microbiology at Aalborg University Hospital. A standard blood culture using two BD BACTEC™ Plus Aerobic medium and one BD BACTEC™ Lytic Anaerobic medium glass culture vials were obtained from a peripheral site upon submission and incubated in the BACTEC FX Top instrument (Becton Dickinson AB, Stockholm, Sweden). The identification of species was conducted with conventional biochemical tests and matrix-assisted laser desorption ionization–time of flight (MALDI Biotyper 3.1, Bruker Daltonics Microflex LT, MBT 6903 MSP Library).

### Plasma preparation and DNA extraction

Plasma was prepared from study blood samples by centrifugation at 1,600g at 4°C for 10 minutes within 1hr from blood draw at the Department of Biochemistry at Aalborg University Hospital. The plasma supernatant was subsequently centrifuged again at 16,000g at 4°C for 10 minutes and the new supernatant was obtained and stored at -80°C until analysis. At analysis, plasma samples were thawed, and single-stranded oligonucleotides were added at a known concentration to allow for absolute quantification of microbial DNA (unpublished). DNA was extracted from 2mL plasma using the QIAamp MinElute ccfDNA kit (Qiagen) standard protocol with DNA elution in 30μL. At least one process control was included in each batch, consisting of 2mL nuclease-free water (Qiagen). DNA was quantified with the Qubit dsDNA HS Assay Kit on a Qubit 4 Fluorometer (Thermo Fisher). DNA length was assessed with the D1000 High Sensitivity ScreenTape Assay on a TapeStation 4150 (Agilent).

### Library preparation and sequencing

10-18μL of extracted DNA was used as input to the SRSLY™ DNA NGS Library Preparation PicoPlus Kit (Claret Biosciences) with custom primers. The library preparation was carried out according to the SRSLY protocol for extreme short fragment retention. At least one process control was included in each batch, consisting of 18μL nuclease-free water (Qiagen). Subsequently, samples were prepared for Oxford Nanopore sequencing with the ligation kit (Oxford Nanopore Technologies, SQK-LSK114). Libraries were sequenced on a PromethION 24 on R10.4.1 flow cells and basecalled with the super-accurate model (Guppy 7.0.9) with a quality cut-off at a phred score of 10 applied.

### Data analysis and bioinformatics

Basecalled reads were demultiplexed, trimmed for adapters, checked for any remaining barcodes or adapters with Cutadapt (version 4.4).^14^ Reads with no alignments to the human genome (BLAST version 2.15.0) were aligned with Bowtie2 to a database consisting of Genome Taxonomy Database (GTDB) representative bacterial and archaeal genomes from release 207 and virus (complete genomes) and fungi (all genomes) from NCBI RefSeq release 215 as well as the reference NCBI Homo sapiens genome (T2T).^15,16^ All identified microbial taxonomies were re-evaluated with proprietary refinement algorithms. Reads from the single-stranded spike-ins were used to quantify DNA for all identified pathogens in genome equivalents per microliter (GPM). The absolute load of microbial DNA in patient samples was compared to the 12 blood donor samples to establish thresholds for clinical relevance.

### Assessment of clinical relevance of mNGS results

To evaluate the relevance of mNGS findings, results were contextualized with the patient’s other microbiological results and overall clinical picture. mNGS results were grouped on a patient level based on the clinical relevance of identifications as adjudicated by two clinical microbiologists (KKS and HLN).^12^ “Confirmed” was assigned only when at least one of the pathogens identified by mNGS was also identified by blood culture. “Probable” was assigned when at least one of the pathogens identified by mNGS was also identified in other samples from the suspected infection site or when the mNGS results aligned very well with the clinical picture (e.g. *Escherichia coli* identified in a patient with suspected urinary tract infection (UTI)). “Possible” was assigned when at least one of the pathogens identified by mNGS was consistent with the clinical picture. “Unlikely” was assigned when the pathogens identified by mNGS were considered irrelevant in the context of the acute infection.

### Potential clinical impact analysis

To evaluate the potential impact on patient antibiotic treatment, the microbial identifications from mNGS were compared to results from routine diagnostics, medical record (suspected site of infection), and the antibiotics administered prior to and during the relevant admission by two clinical microbiologists (KKS and HLN). As the mNGS analysis was conducted in batches following completed patient inclusion, the impact analysis was based on the turnaround time in a potential real-time setup, which was estimated to be around 6 hours (Figure 2b). This aligns with previously reported turnaround times for Nanopore mNGS.^17^

**Figure 1.**
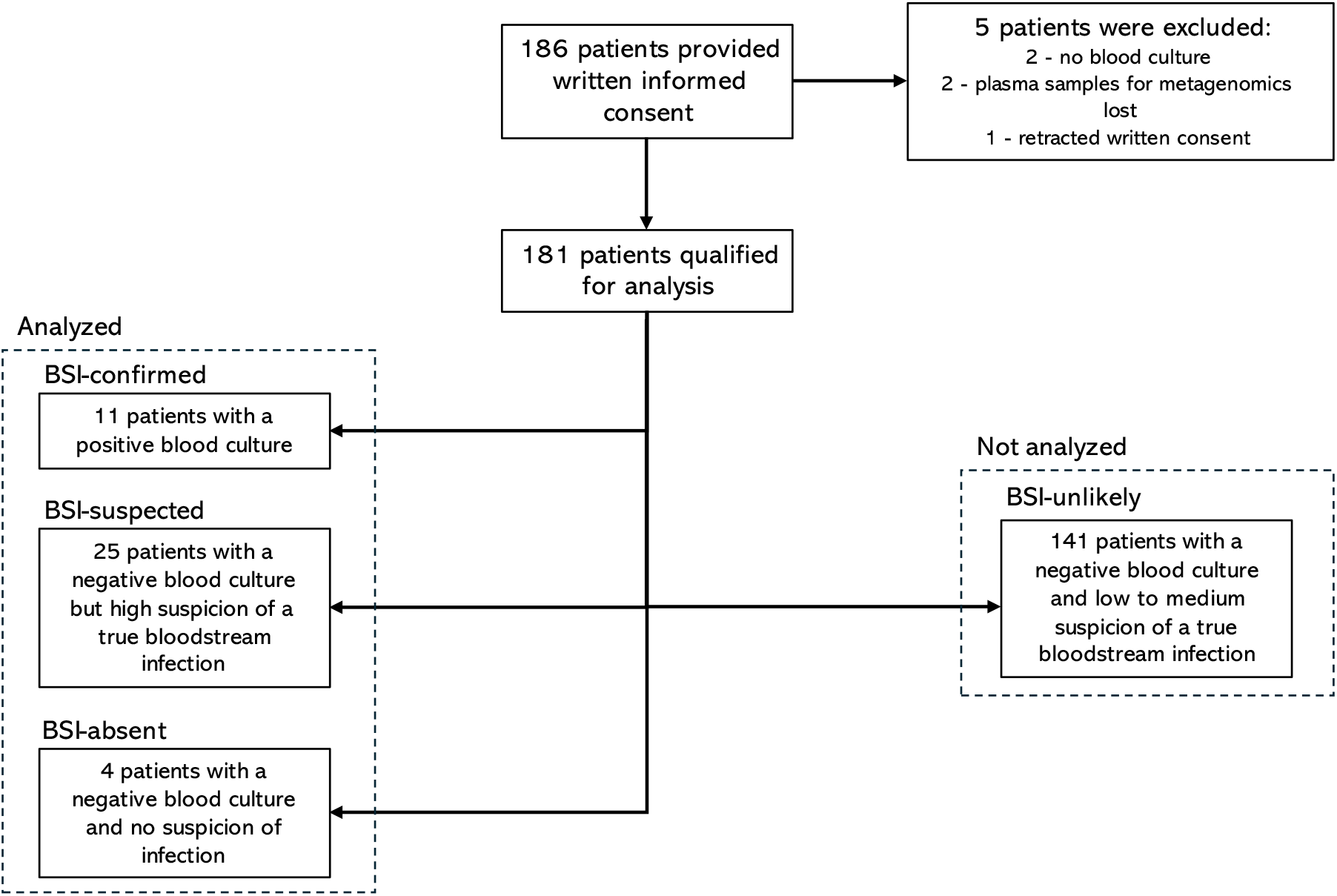
Flowchart of patient inclusion and selection for analysis.

**Figure 2.**
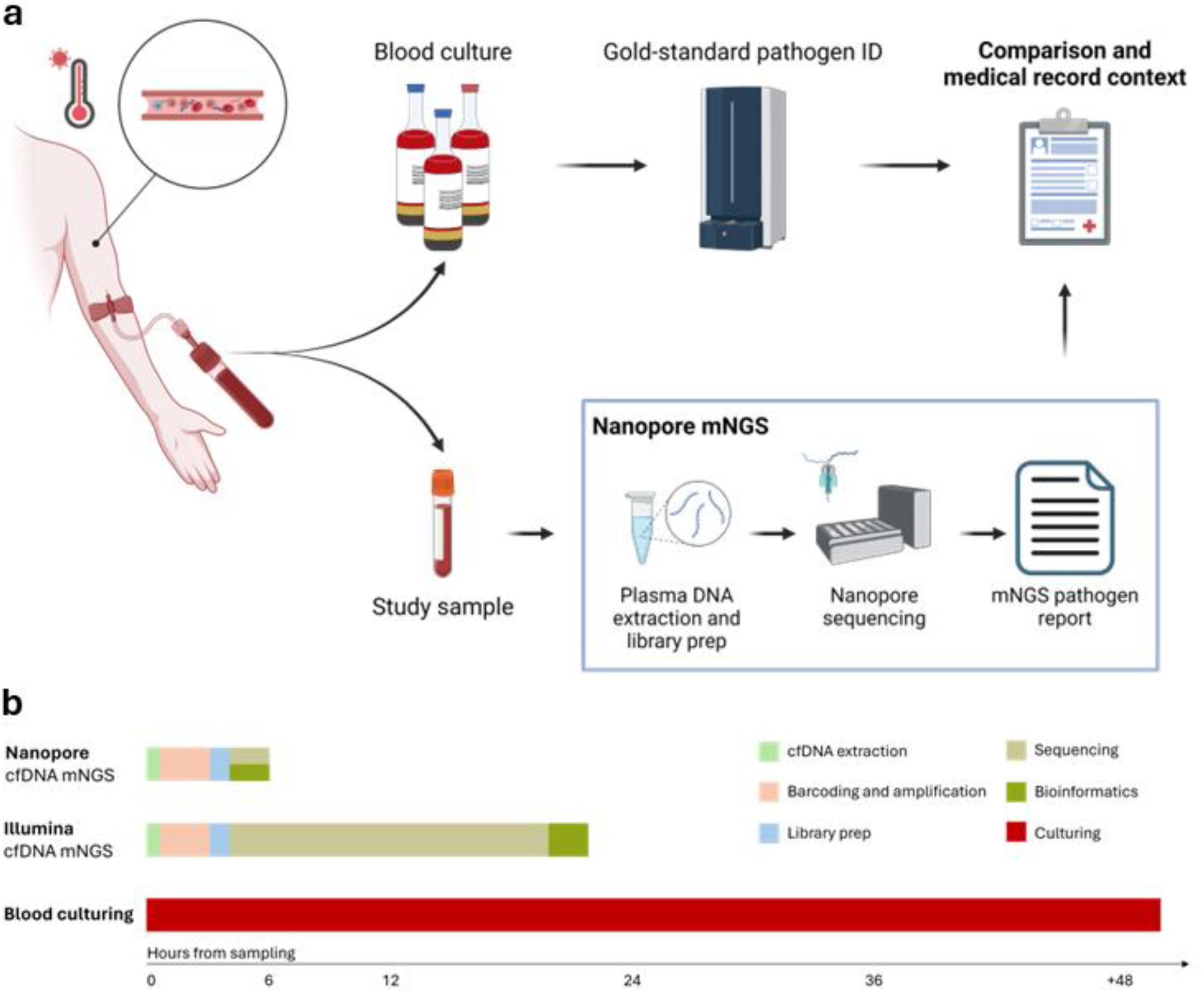
a) Flowchart of the sample handling workflow for blood cultures and mNGS samples (created with biorender.com). b) A timeline for BSI diagnostics with Nanopore mNGS,^17^ Illumina mNGS,^13^ and blood culturing.^7^

### Statistical analysis

Confidence intervals for sensitivity and specificity were calculated using the Clopper-Pearson exact method. If not specified otherwise, a two-sided Wilcoxon-Mann-Whitney test was used to compare distributions. In case of multiple comparisons, the p-values were adjusted with the Holm-Bonferoni method. All statistical analyses were conducted in R (version 4.2.0).

### Ethics

The study protocol was reviewed and accepted by The Scientific Ethics Committee for the North Denmark Region (N-20220015). The study was also approved according to GDPR (article 30) regulation for the North Denmark Region (Project-ID: F2023-056).

### Role of the funding source

The funders of the study had no role in study design, patient inclusion, data analysis, data interpretation, or writing of the manuscript.

## Results

### Patient inclusion and sample selection

During the study period, a total of 186 patients provided written informed consent. Of these, 181 qualified for analysis, and 40 patients were selected after subdividing the patients into four groups: BSI-confirmed (n=11), BSI-suspected (n=25), BSI-absent (n=4), and BSI-unlikely (n=141) (Figure 1). Only samples from patients in BSI-confirmed, BSI-suspected, and BSI-absent were subjected to mNGS analysis.

### Patient characteristics

A subset of 40 patients (50% female) with a median age of 71 years (interquartile range (IQR): 59-84) were analyzed with mNGS (Table 1). Urinary tract infection (UTI, n=12) and lower respiratory tract infection (LRI, n=10) were the most frequent types of infection among the patients. Of the 12 patients admitted with a UTI, six were suspected to have pyelonephritis (Table 2). More than half of the patients had one or more comorbidities, including diabetes (n=13), cancer (n=7), and chronic obstructive pulmonary disease (n=5). Emphasizing that our categorization of BSI was successful, we found that compared to BSI-absent (n=4, median: 1·3 mg/L) and BSI-unlikely (n=141, median: 67 mg/L), the CRP level was significantly higher in patients from BSI-confirmed (n=11, median: 216 mg/L, p=0·047 and p=0·002, Wilcoxon-Mann-Whitney) and BSI-suspected (n=25, median: 204 mg/L, p=0·035 and p<0·0001, Wilcoxon-Mann-Whitney) (appendix p 3).

**Table 1.** Patient characteristics. ^a^Immunosuppresive therapy covers ATC codes L04 and H02. ^2^Oral antibiotics administered at the general practitioner in relation to the relevant infection. ^3^qSOFA, Quick Sequential Organ Failure Assessment.^18^

|  | Patient subgroups |  |  |
| --- | --- | --- | --- |
|  | BSI-confirmed (N=11) | BSI-suspected (N=25) | BSI-absent (N=4) |
| **Patient demographics** |  |  |  |
| Age (years) |  |  |  |
| Median (IQR) | 72 (68·5-81·5) | 67 (59-84) | 56·5 (50·5-67·5) |
| Range | 24-92 | 23-93 | 46-87 |
| Female, n (%) | 3 (27) | 15 (60) | 2 (50) |
| **Comorbidities, n (%)** |  |  |  |
| Any | 4 (36) | 15 (60) | 3 (75) |
| Diabetes | 3 (27) | 9 (36) | 1 (25) |
| Cancer | 1 (9) | 5 (20) | 1 (25) |
| Chronic obstructive pulmonary disease | 1 (9) | 3 (12) | 1 (25) |
| Alcoholism | 1 (9) | 1 (4) | 1 (25) |
| Dialysis | 0 (0) | 0 (0) | 0 (0) |
| **Immunosuppressive therapy<sup>a</sup>, n (%)** | 0 (0) | 2 (8) | 0 (0) |
| **Clinical presentation, n (%)** |  |  |  |
| Oral antibiotics at admission<sup>b</sup> | 0 (0) | 7 (28) | 0 (0) |
| Sepsis according to qSOFA<sup>c</sup> criteria (≥2) | 2 (18) | 4 (16) | 0 (0) |
| **Primary site of infection** |  |  |  |
| Urinary tract infection | 6 (55) | 6 (24) | - |
| Respiratory tract infection | 0 (0) | 10 (40) | - |
| Skin and soft tissue infections | 1 (0) | 4 (8) | - |
| Diverticulitis | 0 (0) | 1 (4) | - |
| PICC-line | 0 (0) | 1 (4) | - |
| Idiopathic pancreatitis | 1 (9) | 0 (0) | - |
| Unknown | 3 (27) | 3 (12) | - |
| 30-day all cause mortality | 0 (0) | 1 (4) | 0 (0) |

**Table 2.**
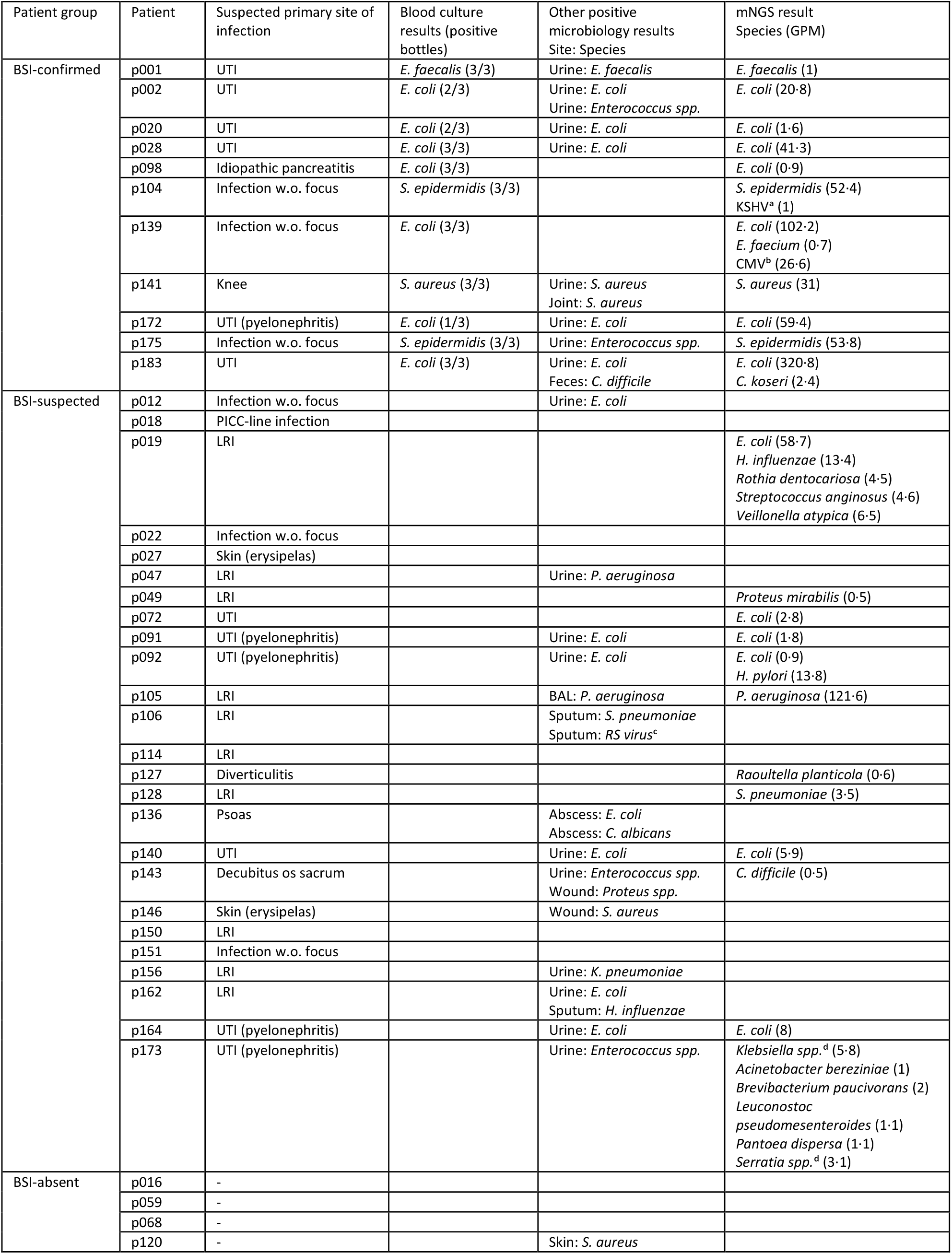
Results from conventional microbiological tests and mNGS. ^a^KSHV: Kaposi’s sarcoma-associated herpesvirus; ^b^CMV: Cytomegalovirus; ^c^RS virus: Human respiratory syncytial virus; ^d^Could not be determined to species level with mNGS.

### Clinical microbiology

The 11 positive blood culture identifications from patients in the BSI-confirmed group comprised *E. coli* (n=7), *Staphylococcus epidermidis* (n=2), *Enterococcus faecalis* (n=1), and *Staphylococcus aureus* (n=1), reflecting that most patients in this group were admitted with a urinary tract focus. Of the 25 patients in BSI-suspected, 14 patients (56%) had positive cultures from urine (n=10), sputum (n=2), bronchoalveolar lavage (n=1), or other sites (n=3), while the remaining patients had no positive microbiological tests to guide the antibiotic treatment. The pathogens identified in other cultures for patients in BSI-suspected were *E. coli* (n=7), *Pseudomonas aeruginosa* (n=2), and others (n=9) (Table 2).

### Metagenomic DNA sequencing analysis

From all 40 patients, plasma cfDNA was sequenced and patient samples had a mean read depth of 10·1M reads (s.d. 5·6M) (appendix p 11). Patients in BSI-confirmed (n=11, median: 10·1 ng/mL) had significantly higher levels of cfDNA in plasma compared to blood donor samples (n=12, median: 2.3 ng/mL) (p=0·00058, Wilcoxon-Mann-Whitney), which was also the case for patients in BSI-suspected (n=25, median: 14·3 ng/mL) (p<0·00001, Wilcoxon-Mann-Whitney) in accordance with previous reports (Figure 3d).^19^

**Figure 3.**
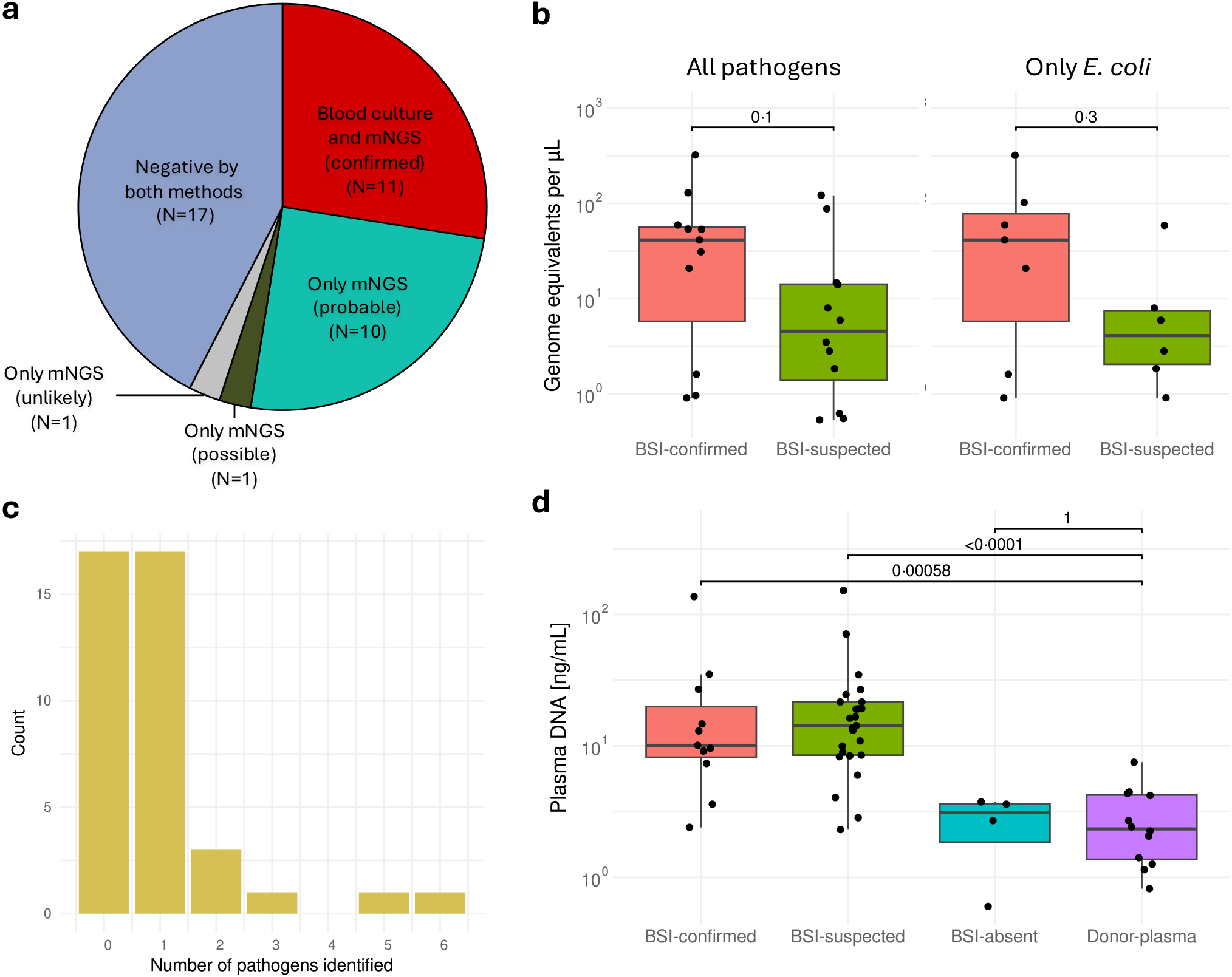
a) Pie chart comparing blood culture and mNGS results. b) Total pathogen burden for patients positive by mNGS in BSI-confirmed and BSI-suspected & compared only for *E. coli* where identified. c) Number of pathogens identified with mNGS per sample. d) cfDNA concentrations across the different patient groups and blood donor samples.

mNGS confirmed all positive blood culture findings, and three patients in this group had 1 or 2 additional pathogens identified (Table 2). The additional findings comprised two bacteria (*Enterococcus faecium* and *Citrobacter koseri*) and two viruses (human cytomegalovirus (human betaherpesvirus 5, CMV) and Kaposi’s sarcoma-associated herpesvirus (human gammaherpesvirus 8, KSHV)). In the BSI-suspected group, 1-6 pathogens were identified in 12 of 25 patients. 11 of the 12 patients with a positive mNGS result had one or more pathogens identified that was deemed relevant for the acute infection by a clinical microbiologist and assigned either ‘Probable’ (n=10) or ‘Possible’ (n=1) (Figure 3a). This included common pathogens like *E. coli, P. aeruginosa*, and *Streptococcus pneumoniae*, but also rarer pathogens like *Serratia spp*., *Raoultella planticola*, and *Veillonella atypica*. The patient with a positive mNGS result, that was categorized as ‘Unlikely’ to be relevant to the acute infection had *Clostridioides difficile* identified. No pathogens were identified in the BSI-absent group. Evaluated against blood culture, the sensitivity was 100% (confidence interval 71·5-100·0%) and the specificity was 59% (confidence interval 38·9-76·5%) on a patient level (Clopper-Pearson exact method).

In five of the 12 patients with a pathogen identified by mNGS, another microbiological test result supported the finding (Table 2). All patients who received antibiotics prior to admission (n=7) were in the BSI-suspected group, of which three (p072, p105, and p140) were positive with mNGS.

With the single-stranded DNA spike-ins the load of pathogen DNA in plasma samples was quantified and pathogens were reported in titers from 0·5-321 genome equivalents per microliter (GPM), which is in accordance with previously reported pathogen DNA titers in BSIs.^20^ Comparing the GPM of *E. coli* - the most frequently identified pathogen - across BSI-confirmed (n=7, median: 41.3 GPM) and BSI-suspected (n=6, median: 4.37 GPM), there was no significant difference (p=0·30, Wilcoxon-Mann-Whitney), which was also the case when comparing the total pathogen load in patients from BSI-confirmed (n=11, median: 41.3 GPM) and BSI-suspected (n=12, median: 4.71 GPM) with a positive mNGS test (p=0·10, Wilcoxon-Mann-Whitney) (Figure 3b).

### Case series

To contextualize the results of mNGS to the patient clinical presentation, we present a more detailed review of selected cases from BSI-confirmed and BSI-suspected groups, who were positive by mNGS. From the BSI-confirmed group three patients were selected for a more detailed analysis.

The first patient, p001, was admitted upon suspicion of a lower respiratory tract infection with a CRP of 109 mg/L (normal reference range <8 mg/L) and treated with empiric IV gentamicin+cefuroxime. During the admission the focus changed to a urinary tract infection with the finding of *E. faecalis* in blood and urine upon which the treatment was changed to ampicillin after 48hrs. mNGS also identified *E. faecalis* at 1 GPM.

The second patient, p104, with known Kaposi’s sarcoma was admitted with acute abdominal pain with a CRP of 41 mg/L and no obvious origin of infection. Treatment was initiated with IV piperacillin+tazobactam and oral ciprofloxacin. 68 hours after admission a blood culture was positive for *S. epidermidis* with the patient’s peripherally inserted central catheter (PICC) line as the suspected origin. This led to a change in treatment to vancomycin and a change of the PICC-line. mNGS identified *S. epidermidis* (52·4 GPM) and Kaposi’s sarcoma-associated herpesvirus (KSHV) (1 GPM).

The third patient, p139, was admitted with infection without focus and elevated CRP of 248 mg/L. The patient had a kidney transplant and received immunosuppressives. *E. coli* was cultured from blood, and this was also identified by mNGS at 102 GPM. Additionally, mNGS identified *Enterococcus faecium* (0·7 GPM) and CMV (26·6 GPM). At day 11 of the hospitalization, a quantitative PCR for CMV was positive with 290 copies/mL.

From the BSI-suspected group three patients were selected for a more detailed description.

The first patient, p128, was admitted upon suspicion of pneumonia with CRP of 470 mg/L and a low systolic blood pressure of 99 mmHg. The patient was known with chronic obstructive pulmonary disease (COPD) and autoimmune hepatitis. The patient was seen by a general practitioner prior to the admission concerning the LRI, but it was unclear from the medical record if the patient was treated with antibiotics at admission. All microbiology tests conducted (blood culture, urine culture, *S. pneumoniae* urine antigen) were negative, while *S. pneumoniae* was identified by mNGS at 3·5 GPM. The patient was administered IV benzylpenicillin at admission with a good clinical response.

The second patient, p173, was admitted upon suspicion of a urinary tract infection and pyelonephritis with a CRP of 164 mg/L. The patient had a positive urine culture with *Enterococcus spp*. as the only positive microbiology result. The mNGS test was positive for several pathogens of which *Klebsiella spp*. were in the highest titer at 5·8 GPM. Three weeks prior to the relevant admission, *K. pneumoniae* was cultured from both blood and urine, suggesting that the mNGS result was relevant.

The third patient, p072, was admitted upon suspicion of a urinary tract infection and pyelonephritis with no positive microbiological tests (blood and urine cultures) during the same admission, while mNGS was positive for *E. coli* (2·8 GPM). At admission the patient was already receiving pivmecillinam ordinated from a general practitioner.

### Potential clinical impact

The mNGS answers for the 22 patients where the result was either confirmed by blood culture or deemed relevant by a clinical microbiologist, were analyzed for potential clinical impact. In a real-time setting, the mNGS result could have affected the antimicrobial treatment in 13 cases (59%) subgrouping to 5 changes from ineffective to effective treatment, 7 escalations from oral to IV, and 1 addition of an extra antimicrobial (more details in appendix p 8). The 5 cases where a change from ineffective to effective treatment could have been facilitated are particularly interesting, and in patients from BSI-confirmed this was achieved by identifying an unsuspected pathogen more rapidly than the blood culture (n=4), while it in patients from BSI-suspected was achieved by identifying an unsuspected pathogen not detected by blood culture (n=1).

## Discussion

In recent years, multiple studies have assessed the use of cfDNA mNGS for BSI diagnostics on patients admitted to e.g. the intensive care unit,^13,21^ the emergency ward,^12^ hematology,^22^ and pediatric departments.^23^ In this study, we show that rapid Nanopore mNGS could be a feasible alternative to blood culturing, as the test confirmed all positive blood cultures (n=11) and provided a relevant pathogen identification in an additional 11 patients. In general, for patients admitted to the emergency ward, no or few microbiological results (e.g. from primary care or previous admissions) are available to guide the upfront antibiotic therapy, and rapid and sensitive infection diagnostics are most welcomed by treating physicians to provide effective antimicrobial therapy for the benefit of the patient and to use narrow-spectrum antibiotics to combat antimicrobial resistance development.^24,25^

The mNGS results reported in this study constitute both common and rare pathogens from various suspected body site origins including UTIs, LRIs, and intraabdominal abscesses, demonstrating applicability across a broad range of etiologies. To the best of our knowledge only a single study with four patients has evaluated the use of Nanopore sequencing for mNGS BSI diagnostics.^26^ Our study establishes the feasibility of using the platform to decrease the turnaround time. We also introduce a quantitative measure of microbial cfDNA in the original sample as a way of standardizing results between patients (and possibly cohorts) and to allow for an improved clinical interpretation of results.

Validation and interpretation of mNGS results in patients with a negative blood culture is difficult and the many additional findings have led to questions on the clinical relevance of these identifications.^27,28^ In the BSI-suspected group from this study, the high number of microbiological tests from other sites that confirm the mNGS result and the fact that patients were included in this group upon suspicion of a systemic infection, supports the relevance of mNGS pathogen identifications. Further, no microbes were identified with mNGS in patients from the BSI-absent group. In addition to the finding of common pathogens, mNGS also identified microbes that were unlikely to be responsible for the acute infection. This includes *H. pylori*, CMV, KSHV, and *C. difficile*, and such findings necessitate that results are interpreted with caution.

We also demonstrate that a fast and sensitive mNGS test has the potential to impact a major fraction of antibiotic treatments. While mNGS tests for clinical microbiology diagnostics are expensive to conduct (the commercially available Karius test is priced at $2000 per sample) the added value of increased sensitivity and fast turnaround time with Nanopore sequencing on antibiotic treatment may impact mortality and length-of-stay to justify this increased cost.^29^ More studies, and randomized interventional studies, are required to answer these questions. We see this test best applied to vulnerable patient groups where etiology determination is crucial for patient survival.

This study has several limitations. Firstly, only patients who could provide informed consent were included and the patients who were in septic shock were thus excluded. Secondly, the study was conducted in a non-real-time setup with batching of samples for analysis after all patients were included in the cohort. Finally, the setup of the study did not allow us to conduct follow-up microbiological tests to investigate mNGS findings.

In summary, this study provides a proof-of-concept for the use of Nanopore cfDNA metagenomics as a diagnostic tool for patients with a BSI. We showed that Nanopore mNGS is capable of identifying a pathogen relevant to the acute infection in twice the number of patients as gold-standard blood culturing, and that the method works on patients with a broad range of etiologies.

## Data Availability

Individual patient metadata is shared in the appendix. Microbial DNA sequencing data from positive mNGS tests is deposited in NCBI SRA and is accessible with the BioProject identifier PRJNA1108520. On reasonable request study protocol and informed consent form, will be shared from the publication date and onwards by contacting the corresponding author.

## Declaration of interests

MEN, SMK, HLN, and MA are in the process of filing patents related to algorithms presented in this study. MEN has stocks in Oxford Nanopore Technologies plc.

## Acknowledgements

The authors would like to thank Kia Brøchner Hjortshøj and Emilia Maria Eriksen Villholth for their efforts to include patients to the study. This research was funded by the Independent Research Fund Denmark (Grant no. 2096-00079B).

## Funding

The study was funded by a research grant from the Independent Research Fund Denmark (Grant no. 2096-00079B).

## Notes

### Summary of Updates

Subheaders in abstract removed as they did not format correctly.

