## appendix for "A new method using rapid Nanopore metagenomic cell-free DNA sequencing to diagnose bloodstream infections: a prospective observational study"

### Rapid metagenomic DNA sequencing to diagnose bloodstream infections in patients admitted to the emergency ward

**Nielsen et al.**

#### Supplementary Methods

##### Potential clinical impact analysis

Patient courses were evaluated for 3 different potential impact types. Firstly, a change from ineffective to effective antibiotic treatment, when DNA sequencing results could have led to a correction from ineffective (either due to antibiotic resistance or the antibiotic not covering the pathogen) to effective antibiotic treatment. This was adjudicated only when the first-line antibiotic against the pathogen identified by DNA sequencing would have been effective. Secondly, an addition of an extra antibiotic for coverage, when DNA sequencing results could have led to the addition of one or more extra antibiotics to cover all identified pathogens relevant to the acute infection. Thirdly, extended treatment, when the finding of a pathogen in the blood with DNA sequencing, that was already covered by the antibiotic treatment could have led doctors to consider prolonging IV antibiotic treatment and admission. Only the pathogens from mNGS adjudicated by the clinical microbiologists as relevant were considered in the impact analysis. In the assessment of impact all of a patient's positive clinical microbiology diagnostics were considered, and e.g. a change to effective treatment was only adjudicated if no relevant diagnostic answers were available at the time (6 hours post admission) to guide the treatment. Negative mNGS answers were not assessed for potential in guiding deescalations.

##### Statistical analysis

The comparison of mNGS results to gold-standard blood culturing was conducted as follows: a true positive was adjudicated when at least one pathogen identified by mNGS was in accordance with blood culturing; a true negative was adjudicated when no pathogens were identified by neither blood culture or mNGS; a false positive was adjudicated when blood culture was negative and mNGS was positive; a false negative was adjudicated when blood culture was positive and mNGS was negative for that pathogen.[2]

If not specified otherwise, a two-sided Wilcoxon-Mann-Whitney test was used to compare distributions. In case of multiple comparisons, the p-values were adjusted with the Holm-Bonferoni method. All statistical analyses were conducted in R (version 4.2.0).

**Supplementary Figures**

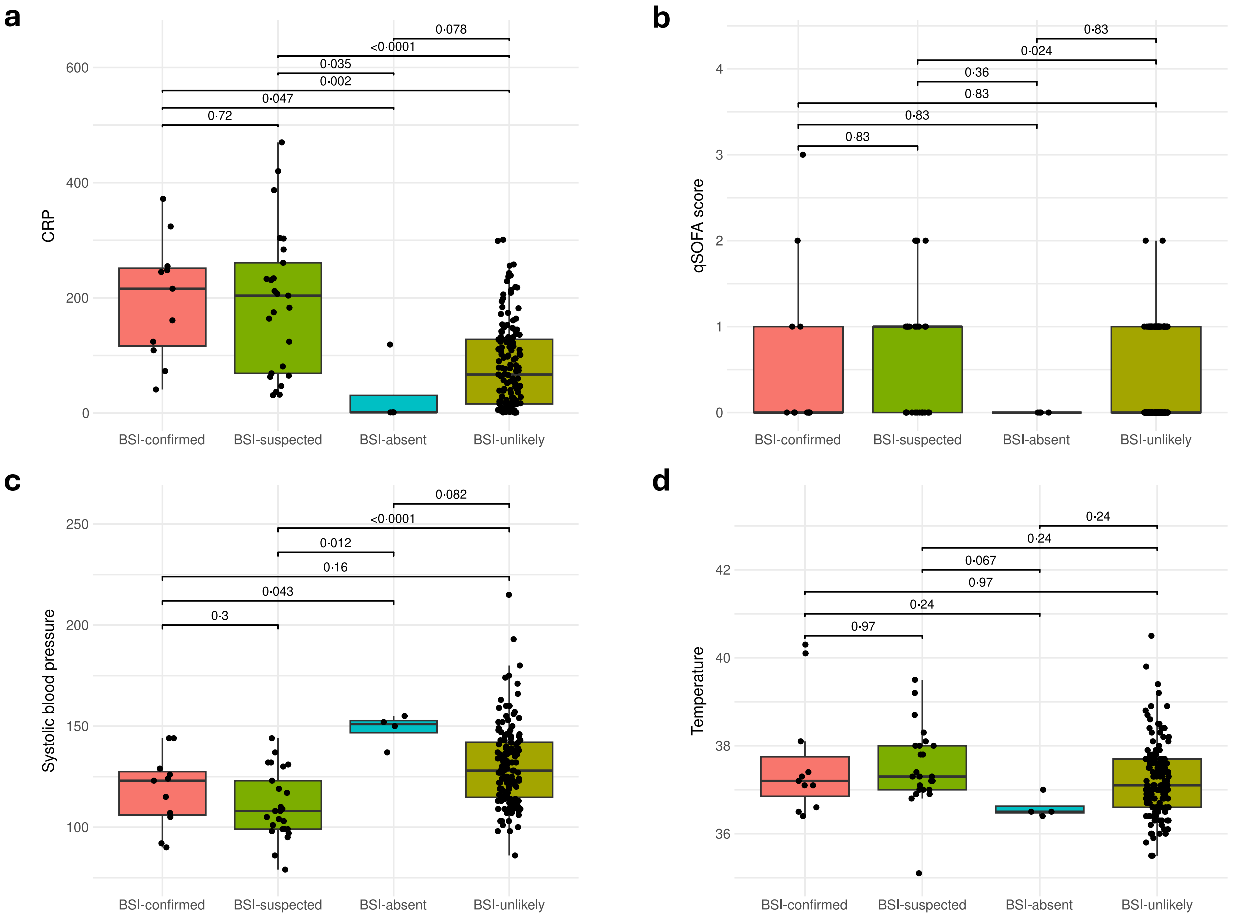

**Figure S1. Clinical characteristics of patients who were analyzed and excluded.**
a) CRP at blood culture for BSI-confirmed, BSI-suspected, BSI-absent, and BSI-unlikely groups. b) qSOFA score at blood culture for the four groups. c) Systolic blood pressure at blood culture for the four groups. d) Temperature at blood culture for the four groups.

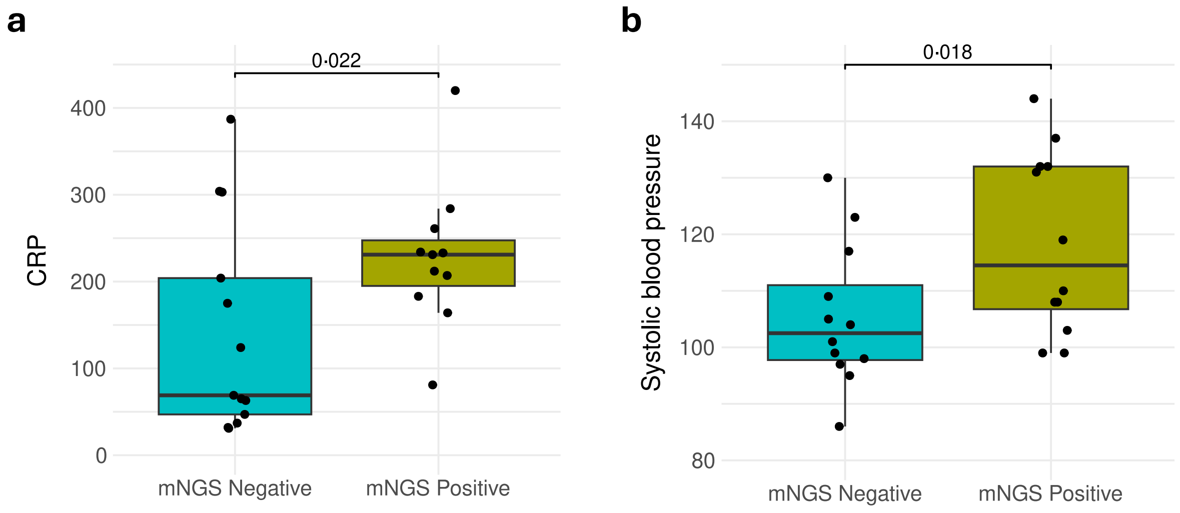

**Figure S2. Clinical characteristics for patients in BSI-suspected with negative and positive mNGS results.** a) CRP at blood culture. b) Systolic blood pressure at blood culture.

**Supplementary Tables**

**Table S1. Detailed metadata on clinical presentation and antibiotics for all patients.**

| Patient | Suspected primary infection site | CRP (mg/L) | Systolic blood pressure (mmHg) | Temperature (°C) | WBC ($10^3$ cells/mm³) | RF (1/min) | GCS | qSOFA | Antibiotics prior to admission | Antibiotics during admission |
|---|---|---|---|---|---|---|---|---|---|---|
| p001 | LRI | 109 | 124 | 40.1 | 10.3 | 30 | 13 | 2 | | Gentamicine \| Cefuroxime \| Benzylpenicillin \| Ampicillin \| Piperacillin/tazobactam |
| p002 | UTI | 216 | 90 | 36.6 | 10.7 | 24 | 14 | 3 | | Gentamicin \| Piperacillin/tazobactam \| Meropenem \| Ciprofloxacin \| Ampicillin |
| p020 | UTI | 161 | 115 | 36.4 | 14.8 | 20 | 15 | 0 | | Piperacillin/tazobactam \| Mecillinam \| Pivmecillinam |
| p028 | LRI | 255 | 105 | 37.1 | 24.8 | 24 | 15 | 1 | | Amoxicillin \| Piperacillin/tazobactam |
| p098 | Idiopathic pancreatitis | 73 | 144 | 37.3 | 14.5 | 16 | 15 | 0 | | Piperacillin/tazobactam \| Metronidazole |
| p104 | Infection w.o. focus | 41 | 123 | 36.5 | 7.5 | 16 | 15 | 0 | | Piperacillin/tazobactam \| Ciprofloxacin \| Vancomycin |
| p139 | Infection w.o. focus | 248 | 129 | 37.4 | 5.3 | 18 | 15 | 0 | | Piperacillin/tazobactam \| Ciprofloxacin |
| p141 | Knee | 324 | 144 | 37.2 | 5.3 | 12 | 15 | 0 | | Cloxacillin \| Dicloxacillin \| Benzylpenicillin \| Cefuroxime |
| p172 | UTI | 245 | 107 | 40.3 | 17.1 | 16 | 15 | 0 | | Gentamicin \| Ampicillin \| Piperacillin/tazobactam \| Pivmecillinam \| Phenoxymethylpenicillin |
| p175 | Infection w.o. focus | 124 | 92 | 38.1 | 13.6 | 19 | 15 | 1 | | Piperacillin/tazobactam \| Vancomycin |
| p183 | UTI | 372 | 126 | 37.1 | 19.4 | 18 | 15 | 0 | | Piperacillin/tazobactam \| Ampicllin \| Metronidazole |
| p012 | Infection w.o. focus | 204 | 86 | 36.8 | 7.6 | 23 | 15 | 2 | Metronidazole | Nystatin \| Amoxicillin \| Piperacillin/tazobactam |
| p018 | PICC-line infection | 37 | 95 | 36.9 | 4.8 | 12 | 15 | 1 | | Benzylpenicillin \| Fluconazole \| Ampicillin \| Anidulafungin |
| p019 | LRI | 284 | 103 | 38 | 10.8 | 22 | 15 | 1 | | Ampicillin \| Gentamicin \| Piperacillin/tazobactam |
| p022 | Infection w.o. focus | 175 | 101 | 35.1 | 18.1 | 16 | 15 | 0 | Phenoxymethyl-penicillin | Ampicillin \| Piperacillin/tazobactam \| Moxifloxacin |

| | | | | | | | | | | |
|---|---|---|---|---|---|---|---|---|---|---|
| p027 | Skin | 63 | 99 | 37.3 | 10.9 | 18 | 15 | 1 | | Fluconazole \| Clarithromycin \| Piperacillin/tazobactam \| Dicloxacillin |
| p047 | LRI | 124 | 130 | 39.2 | 23.3 | 22 | 15 | 1 | | Benzylpenicillin \| Piperacillin/tazobactam \| Amoxicillin |
| p049 | LRI | 420 | 131 | 37.3 | 27.7 | 16 | 15 | 0 | | Cefuroxime \| Clarithromycin \| Roxithromycin \| Moxifloxacin |
| p072 | UTI | 207 | 144 | 37 | 15.5 | 16 | 15 | 0 | Pivmecillinam | Ampicillin \| Pivampicillin \| Pivmecillinam \| Gentamicin |
| p091 | UTI | 261 | 108 | 37.2 | 12.3 | 14 | 15 | 0 | | Piperacillin/tazobactam \| Pivmecillinam |
| p092 | UTI | 212 | 108 | 37 | 10 | 14 | 15 | 0 | | Piperacillin/tazobactam \| Pivmecillinam |
| p105 | LRI | 233 | 137 | 38.7 | 23.4 | 22 | 15 | 1 | Azithromycin | Clarithromycin \| Benzylpenicillin \| Piperacillin/tazobactam \| Amoxicillin \| Moxifloxacin |
| p106 | LRI | 387 | 117 | 37 | 13.1 | 22 | 15 | 1 | Unknown antibiotic | Piperacillin/tazobactam \| Phenoxymethylpenicillin |
| p114 | LRI | 69 | 97 | 37.2 | 12.3 | 22 | 15 | 2 | | Benzylpenicillin \| Amoxicillin |
| p127 | Diverticulitis | 183 | 99 | 37.1 | 16.3 | 22 | 15 | 2 | | Metronidazole \| Benzylpenicillin \| Gentamicin \| Amoxicillin |
| p128 | LRI | 470 | 99 | 36.9 | 12.2 | 20 | 15 | 1 | | Benzylpenicillin \| Phenoxymethylpenicillin |
| p136 | Psoas | 303 | 105 | 37.8 | 15.4 | 12 | 15 | 0 | Unknown antibiotic | Metronidazole \| Piperacillin/tazobactam \| Fluconazole |
| p140 | UTI | 81 | 132 | 38 | 13.6 | 16 | 15 | 0 | Pivmecillinam | Ampicillin \| Gentamicin \| Pivmecillinam |
| p143 | Decubitus os sacrum | 231 | 110 | 37.8 | 14.1 | 18 | 14 | 1 | | Cefuroxime \| Amoxicillin |
| p146 | Skin | 65 | 104 | 36.9 | 10.5 | 18 | 15 | 0 | | Benzylpenicillin \| Cloxacillin \| Phenoxymethylpenicillin |
| p150 | LRI | 31 | 109 | 39.5 | 1.4 | 18 | 15 | 0 | | Ciprofloxacin \| Piperacillin/tazobactam \| Gentamicin \| Meropenem |
| p151 | Infection w.o. focus | 304 | 123 | 38.1 | 13 | 16 | 15 | 0 | | Cefuroxime \| Trimethoprim |
| p156 | LRI | 47 | 79 | 37.4 | 28.9 | 20 | 15 | 1 | | Piperacillin/tazobactam \| Amoxicillin |
| p162 | LRI | 32 | 98 | 37.3 | 9.4 | 28 | 15 | 2 | | Meropenem \| Ciprofloxacin |

| | | | | | | | | | | |
|---|---|---|---|---|---|---|---|---|---|---|
| p164 | UTI | 234 | 119 | 38.3 | 7.3 | 14 | 15 | 0 | | Piperacillin/tazobactam \| Ciprofloxacin |
| p173 | UTI | 164 | 132 | 38 | 10 | 14 | 15 | 0 | | Vancomycin \| Pivmecillinam \| Ciprofloxacin |
| p016 | | 119 | 152 | 36.5 | 9.8 | 14 | 15 | 0 | | |
| p059 | | 1.2 | 155 | 36.4 | 6.4 | 20 | 15 | 0 | | |
| p068 | | 1.3 | 150 | 36.5 | 10.1 | 16 | 15 | 0 | | |
| p120 | | 1.3 | 137 | 37 | 9.6 | 16 | 15 | 0 | | Dicloxacillin |

Abbreviations: RF = Respiratory frequency, WBC = White blood cell count, CRP = C-reactive protein, qSOFA = Quick Sequential Organ Failure Assessment,[3] GCS = Glasgow Coma Scale.

**Table S2. Detailed metadata on the assessment of relevance of mNGS findings.**

| Patient | Blood culture result | Secondary microbiology result | mNGS result (GPM) | Clinical relevance of mNGS result | Potential impact on antibiotic treatment | Comments related to treatment impact |
|---|---|---|---|---|---|---|
| p001 | *Enterococcus faecalis* | Urine: *E. faecalis* | *E. faecalis* (1) | Confirmed | Change | From gentamicin and cefuroxime to ampicillin |
| p002 | *Escherichia coli* | Urine: *E. coli*, Urine: *Enterococcus* spp. | *E. coli* (20·8) | Confirmed | None | |
| p020 | *E. coli* | Urine: *E. coli* | *E. coli* (1·6) | Confirmed | None | |
| p028 | *E. coli* | Urine: *E. coli* | *E. coli* (41·3) | Confirmed | None | |
| p098 | *E. coli* | | *E. coli* (0·9) | Confirmed | None | |
| p104 | *Staphylococcus epidermidis* | | *S. epidermidis* (52·4)<br>KSHV (1) | Confirmed | Change | From piperacillin/tazobactam to vancomycin |
| p139 | *E. coli* | | *E. coli* (102·2)<br>*Enterococcus faecium* (0·7)<br>CMV (26·6) | Confirmed | Addition | Addition of antiviral treatment |
| p141 | *Staphylococcus aureus* | Urine: *S. aureus*, Joint: *S. aureus* | *S. aureus* (31) | Confirmed | Change | Patient was discharged without any antibiotic treatment and readmitted and started cloxacillin the next day when the blood culture was positive. |
| p172 | *E. coli* | Urine: *E. coli* | *E. coli* (59·4) | Confirmed | None | |
| p175 | *S. epidermidis* | Urine: *Enterococcus spp.* | *S. epidermidis* (53·8) | Confirmed | Change | From piperacillin/tazobactam to vancomycin |
| p183 | *E. coli* | Urine: *E. coli*, Feces: *Clostridiodes difficile* | *E. coli* (320·8)<br>*Citrobacter koseri* (2·4) | Confirmed | None | |

| | | | | | | |
|---|---|---|---|---|---|---|
| p019 | | | *E. coli* (58·7)<br>*Haemophilus influenzae* (13·4)<br>*Rothia dentocariosa* (4·5)<br>*Streptococcus anginosus* (4·6)<br>*Veillonella atypica* (6·5) | Probable | None | |
| p049 | | | *Proteus mirabilis* (0·5) | Possible[a] | None | |
| p072 | | | *E. coli* (2·8) | Probable | Escalation | Extended duration of IV antibiotic treatment |
| p091 | | Urine: *E. coli* | *E. coli* (1·8) | Probable | Escalation | Extended duration of IV antibiotic treatment |
| p092 | | Urine: *E. coli* | *E. coli* (0·9)<br>*H. pylori* (13·8) | Probable | None | |
| p105 | | BAL: *Pseudomonas aeruginosa* | *P. aeruginosa* (121·6) | Probable | Change | From penicillin and clarithromycin to piperacillin/tazobacta m and ciprofloxacin |
| p127 | | | *Raoultella planticola* (0·6) | Probable | Escalation | Peroral escalation from amoxicillin to ciprofloxacin |
| p128 | | | *S. pneumoniae* (3·5) | Probable | Escalation | Extended duration of IV antibiotic treatment |
| p140 | | Urine: *E. coli* | *E. coli* (5·9) | Probable | Escalation | Extended duration of IV antibiotic treatment |
| p143 | | Urine: *Enterococcus spp.*, Wound: *Proteus spp.* | *C. difficile* (0·5) | Unlikely | None | |
| p164 | | Urine: *E. coli* | *E. coli* (8) | Probable | Escalation | Extended duration of IV antibiotic treatment |
| p173 | | Urine: *Enterococcus spp.* | *Klebsiella spp.* (5·8)<br>*Acinetobacter bereziniae* (1)<br>*Brevibacterium paucivorans* (2)<br>*Leuconostoc pseudomesenteroides* (1·1) | Probable | Escalation | Extended duration of IV antibiotic treatment |

| | | | |
|---|---|---|---|
| | | | *Pantoea dispersa* (1·1)<br>*Serratia spp.* (3·1) |

Abbreviations: GPM = Genome Equivalents per microliter, KSHV = Kaposi's sarcoma-associated herpesvirus, CMV = Cytomegalovirus, BAL = bronchoalveolar lavage

[a]Consistent with a bronchial lavage culture from a previous hospital admission, which supports the finding of *P. mirabilis* in a patient with LRI focus.

**Table S3. Read depth and plasma cfDNA concentration for all patients and blood donors.**

| Patient | DNA concentration (ng/mL) | Read depth (M) |
|---|---|---|
| p001 | 9·6 | 2·69 |
| p002 | 3·6 | 2·1 |
| p012 | 14·3 | 14·2 |
| p016 | 3·8 | 5·72 |
| p018 | 9·0 | 5·22 |
| p019 | 70·9 | 10·2 |
| p020 | 14·7 | 1·15 |
| p022 | 4·1 | 8·11 |
| p027 | 13·1 | 18·7 |
| p028 | 27·0 | 9·15 |
| p047 | 24·6 | 21·4 |
| p049 | 19·2 | 5·94 |
| p059 | 3·6 | 9·94 |
| p068 | 2·7 | 0·629 |
| p072 | 19·1 | 7·18 |
| p091 | 8·4 | 10·3 |
| p092 | 8·5 | 5·44 |
| p098 | 7·4 | 9·99 |
| p104 | 2·4 | 16·9 |
| p105 | 21·6 | 5·61 |
| p106 | 26·9 | 6·48 |
| p114 | 6·0 | 18·2 |
| p120 | 0·6 | 1·67 |
| p127 | 2·8 | 11·8 |
| p128 | 34·8 | 17·3 |
| p136 | 16·6 | 18·3 |
| p139 | 10·1 | 9·4 |

| | | |
|---|---|---|
| p140 | 10·0 | 16·6 |
| p141 | 35·1 | 5·65 |
| p143 | 19·2 | 11·3 |
| p146 | 2·3 | 11·7 |
| p150 | 152·1 | 6·3 |
| p151 | 16·3 | 11·6 |
| p156 | 10·9 | 4·22 |
| p162 | 8·3 | 12·7 |
| p164 | 21·6 | 12·6 |
| p172 | 13·0 | 15·5 |
| p173 | 13·7 | 10·4 |
| p175 | 9·1 | 21 |
| p183 | 136·8 | 10·5 |
| d001 | 4·3 | 6·47 |
| d002 | 4·5 | 10·3 |
| d003 | 2·1 | 8·57 |
| d004 | 1·3 | 10·4 |
| d005 | 1·1 | 5·48 |
| d006 | 2·4 | 13·6 |
| d007 | 1·4 | 3·24 |
| d008 | 0·8 | 14·5 |
| d009 | 7·5 | 11·6 |
| d010 | 2·3 | 16·7 |
| d011 | 4·2 | 17·6 |
| d012 | 2·7 | 2 |
